# Modeling the Impact of Social Determinants on Breast Cancer Screening: A Data-Driven Approach

**DOI:** 10.1101/2025.06.24.25330138

**Authors:** Guofang Ma, Miranda G. Scully, Jiahui Luo, Wesley J. Marrero, Jiazuo H. Feng, Christine M. Gunn, Roberta M. diFlorio-Alexander, Anna N.A. Tosteson, Sally A. Kraft

## Abstract

**Background:** This study addresses the critical implementation science challenge of operationalizing social determinants of health (SDoH) in clinical practice. We develop and validate models demonstrating how SDoH predicts mammogram screening behavior within a rural population. Our work provides healthcare systems with an evidence-based framework for translating SDoH data into effective interventions.

**Methods:** We model the relationship between SDoH and breast cancer screening adherence using data from over 63,000 patients with established primary care relationships within the Dartmouth Health System. Our analytical framework integrates multiple machine learning techniques including light gradient boosting machine, random forest, elastic-net logistic regression, Bayesian regression, and decision tree classifier with SDoH questionnaire responses, demographic information, geographic indicators, insurance status, and clinical measures to quantify and characterize the influence of SDoH on mammogram scheduling and attendance.

**Results:** Our models achieve moderate discriminative performance in predicting screening behaviors, with an average area under the receiver operating characteristic curve (ROC AUC) of 71% for scheduling and 70% for attendance in validation datasets. Key social factors influencing screening behaviors include geographic accessibility measured by the rural-urban commuting area, neighborhood socioeconomic status captured by the area deprivation index, and healthcare access factors related to clinical sites. Additional influential variables include months since the last mammogram, current age, and the Charlson comorbidity score, which intersect with social factors influencing healthcare utilization. By systematically modeling these SDoH and related factors, we identify opportunities for healthcare organizations to transform SDoH data into targeted, facility-level intervention strategies while adapting to payer incentives and addressing screening disparities.

**Conclusions:** Our model provides healthcare systems with a data-driven approach to understanding and addressing how SDoH shape mammogram screening behaviors, particularly among rural populations. While initially focused on breast cancer screening, this systematic framework lays the groundwork for analyzing SDoH’s influence on other preventive health behaviors, demonstrating the potential for broader applications in improving routine preventive care utilization.

## 1. INTRODUCTION

The integration of social determinants of health (SDoH) into clinical practice has emerged as a vital frontier in healthcare delivery transformation. Healthcare systems increasingly recognize that addressing SDoH can significantly impact health outcomes and costs (Mohan and Gaskin, 2024). Recent evidence demonstrates that higher SDoH needs correlate with greater expenses across both commercial and public insurance systems (Mohan and Gaskin, 2024). This recognition has highlighted the need for financial incentives for healthcare organizations to incorporate SDoH data into their clinical workflows and decision-making processes(Rangachari and Thapa, 2025). Within this evolving landscape, breast cancer screening provides an ideal context for examining SDoH integration, as mammography represents a preventive service with well-documented benefits (US Preventive Services Task Force, 2024). However, despite being an effective early detection tool for breast cancer—the second leading cause of cancer-related deaths among women globally—mammography screening rates consistently fall below national targets (Pace and Keating, 2024). This gap between evidence-based recommendations and practice points reflects underlying barriers that extend beyond clinical factors alone.

Various obstacles to breast cancer screening adherence have been previously documented, including socio-economic challenges (Miller *et al*., 2019; Ponce-Chazarri *et al*., 2023), insurance status (Miller *et al*., 2019), geographic accessibility (Miller *et al*., 2019; Pohl *et al*., 2025), transportation limitations (Pohl *et al*., 2025), cultural beliefs (Lofters *et al*., 2017; Albadawi, Alsharawneh and Othman, 2025), health literacy levels (Ponce-Chazarri *et al*., 2023), and provider communication effectives (Albadawi, Alsharawneh and Othman, 2025). Collectively, these studies illustrate how personal, social, and systemic factors can intertwine to create complex patterns of healthcare utilization and screening behaviors (Coughlin, 2019). Understanding these patterns requires recognizing that social determinants do not operate in isolation but rather form inter-connected networks of influence that shape individual health decisions.

While the relationships between SDoH and screening behaviors are well-documented, operationalizing SDoH data to improve screening outcomes still presents significant methodological challenges (Andermann, 2016; Novilla *et al*., 2023; Ganatra *et al*., 2024). The intricate connections between various social determinants and their variable impacts on clinical outcomes demand sophisticated analytical approaches beyond traditional methods. Qualitative research has provided valuable foundations for identifying the multi-faceted nature of social factors influencing screening behaviors. For example, prior work has explored how economic stability and healthcare access barriers shape lung cancer screening decisions among Latino communities (Alaniz-Cantú *et al*., 2024), how health system organizational factors create barriers to implementing social needs screening in primary care settings (Kazi *et al*., 2024), and how geographic and socioeconomic factors influence cancer care trajectories and access to treatment (Teteh *et al*., 2023). However, these qualitative studies are inherently limited in their ability to analyze the complex interactions among these factors at scale.

Machine learning approaches offer promising solutions to this complexity, enabling healthcare systems to analyze patterns within SDoH data and develop targeted interventions. These analytical techniques can identify subtle relationships across multiple social determinants simultaneously, which helps to reveal insights that might remain obscured using conventional methods. By applying machine learning to SDoH data in the context of breast cancer screening, healthcare organizations can develop personalized approaches to improving screening rates and meet their adherence targets.

In striving towards operationalizing SDoH data and overcoming the limitations of traditional analytical approaches, our study presented an integrated approach to predicting breast cancer screening behaviors. We first developed a generalizable framework for modeling the relationships between social determinants and mammogram scheduling and attendance, providing a structured approach to quantifying these complex influences. We then applied machine learning techniques to transform SDoH data into actionable insights that healthcare systems can use to improve mammogram adherence rates. Through this integrated approach, we aimed to create an evidence-informed methodology for leveraging SDoH data to enhance breast cancer screening outcomes while providing a replicable model that organizations can adapt for other preventive services and health outcomes. This work contributed to the implementation science pipeline by providing healthcare systems with quantitative tools to systematically translate SDoH data into actionable screening interventions.

## 2. MATERIALS AND METHODS

### 2.1 General Framework for SDoH Analysis in Mammogram Screening Behavior

Our generalizable framework included the following steps: data pre-processing and variable construction, model selection and implementation, performance evaluation, and model explainability analysis. We detailed these steps and presented their execution for predicting the probability of mammogram screening behaviors, including both scheduling and attendance.

#### 2.1.1 Data Pre-processing and Variable Construction

Our analytical process began by preprocessing three categories of predictor variable that shaped mammogram screening behavior: SDoH, demographic characteristics, and geographic factors, along with defining our outcome variables (scheduling and attendance status). We established a threshold for variable inclusion based on data completeness, excluding variables with excessive missingness (>80%). While higher than conventional thresholds, this approach was necessary to retain sufficient SDoH variables for analysis, given the inherently sparse nature of such data in clinical practice. We addressed potential concerns about data quality through our imputation strategy and sensitivity analyses using complete case analysis (Section 2.3). For standardization, we standardized SDoH questionnaire variables by merging duplicate questions that assessed similar constructs, retaining the question with more complete responses and consolidating the data. We also converted inconsistent missing value representations including blank entries, ‘N/A’, ‘NULL’, and ‘Unknown’ responses to standardized missing values, while preserving meaningful response categories such as ‘Patient Refused’ and ‘Choose not to disclose’ as distinct categorical levels for analysis.

To facilitate a robust analysis of our demographic information, we consolidated language preferences into appropriate categories according to data availability (e.g., English and Others). We also combined race and ethnicity into major categories based on available data, and merged insurance type into primary categories that reflected common coverage patterns. For geographic variables, we processed the rural-urban commuting area (RUCA) code and area deprivation index (ADI) to capture spatial dimensions of healthcare access and quantify neighborhood socio-economic status. We removed the ADI national rank variable and retained only the ADI state rank for analysis. We also converted problematic ADI state rank codes including ‘GQ’ (Group Quarters), ‘PH’ (Other US territories), and ‘QDI’ (Quality Data Issues) to missing values to ensure our data quality.

Our framework assumed a binary outcome indicating whether a patient had scheduled a screening (1) or not (0), and another indicating whether a patient attended their scheduled mammogram (1) or not (0). For the attendance framework, we only included patients who had a mammogram scheduled, determining attendance based on whether they received a mammogram within the past 27 months. This timeframe aligned with Dartmouth Health’s internal clinical guidelines, which recommend women ages 50-75 with average risk receive mammogram screening every two years at a minimum, with a 27-month compliance window to account for scheduling delays. For the scheduling model, we included months since the last mammogram as a predictor variable; however, for the attendance model, we excluded this variable to avoid redundancy and potential data leakage since the 27-month timeframe was already incorporated into the attendance outcome definition. These screening statuses served as a direct indicator of patient engagement with screening guidelines and acted as the target variables for our predictive models.

For both our scheduling and attendance analyses, we employed data imputation techniques using the missForest algorithm (Stekhoven and Bühlmann, 2012), a non-parametric imputation method based on random forests that can handle mixed-type data and complex interactions. This approach was necessary because relying solely on complete records would have substantially reduced sample sizes, which might have limited the robustness and generalizability of our models. We applied missForest separately to stratified training and testing sets to prevent data leakage, with imputation parameters including mtry set to the square root of the number of variables and parallel processing to enhance computational efficiency. This imputation strategy enabled us to utilize information from a much larger patient population while addressing missing data in a principled way.

#### 2.1.2 Model Comparison

Our framework compared the performance of machine learning approaches for predicting a binary outcome of interest. For predicting mammogram scheduling, we compared elastic-net logistic regression (Zou and Hastie, 2005), random forest (Breiman, 2001), and light gradient boosting machine (Ke *et al*., 2017). For predicting attendance, we selected different algorithms due to the smaller expected sample size (only patients who had scheduled mammograms), comparing Bayesian regression with prior log-odds based on previous literature (Gelman *et al*., 2008), elastic-net logistic regression (Zou and Hastie, 2005), and decision tree classifier (Breiman, 2001). These machine learning techniques provided analytical strengths while maintaining interpretability for healthcare practitioners.

#### 2.1.3 Performance Evaluation

We developed a nested cross-validation scheme that incorporated 10 outer folds for model assessment and 5 inner folds for parameter optimization (Figure 1). This approach systematically tested how well our models would perform when applied to new patients not used in model development. By repeatedly training and testing on different patient subsets, this method provided a realistic estimate of model performance for healthcare systems implementing these tools in clinical practice.

**Figure 1:**
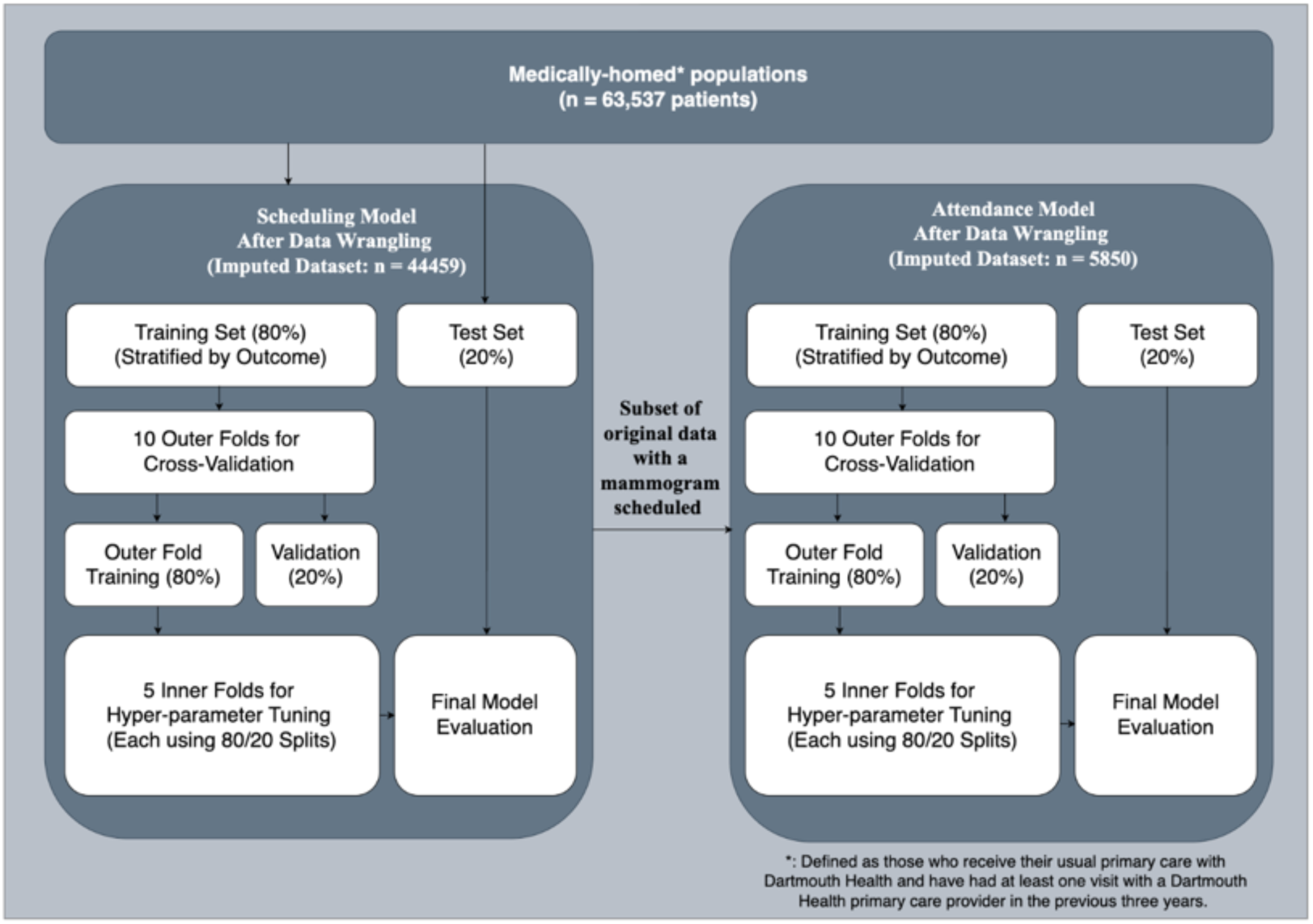
Data Splitting Process for both Scheduling and Attendance Model (Base Analysis with Imputed Data).

The data splitting process followed a stratified approach: we first divided the data into training (80%) and testing (20%) sets, then further divided the outer training data into inner training (80%) and validation (20%) subsets iteratively while maintaining the original distribution of mammogram scheduling behaviors (Varma and Simon, 2006). This structure allowed healthcare systems to assess how well the models might generalize across different patient populations and implementation contexts.

Our performance assessment utilized the Area Under the Receiver Operating Characteristic curve (AUC-ROC) as the primary metric, chosen for its ability to evaluate model discrimination across different contexts (Sohil, Sohali and Shabbir, 2022). To address class imbalance challenges common in screening programs, we implemented the Random Over-Sampling Examples (ROSE) technique with synthetic information, ensuring our models effectively learned from both screened and unscreened populations (Lunardon, Menardi and Torelli, 2014).

To assess the statistical significance of performance differences between models, we conducted pairwise comparisons using DeLong’s test for correlated ROC curves (DeLong, DeLong and Clarke-Pearson, 1988).

#### 2.1.4 Model Explainability Analysis

For the best-performing models, we conducted permutation importance analysis to measure the relative contribution of each predictor to the probability of mammogram scheduling and attendance (Altmann *et al*., 2010). This analysis provided actionable insights for implementation strategies, measuring the relative contribution of each social driver to predicting non-screening behavior. The relationships between key predictors and screening behaviors were further visualized through partial dependence plots (Friedman, 2001), providing healthcare practitioners with intuitive tools for understanding how SDoH influenced screening behaviors in their specific implementation context.

### 2.2 Case Study: The Dartmouth Health System

While the framework is designed to be generalizable across different healthcare systems, we applied it specifically to the Dartmouth Health System to demonstrate its practical utility and effectiveness in a real-world setting. Dartmouth Health is an academic health system serving patients across northern New England through seven hospitals and affiliated ambulatory clinics (Dartmouth Health, 2025). It includes the academic campus and community and critical access hospitals in Vermont and New Hampshire. With over 16,000 employees including 2,300 providers, the system delivers approximately 3 million outpatient visits annually and is recognized as a nationwide leader in rural health (Dartmouth Health, 2025). This application allowed us to assess the framework’s ability to generate actionable insights within a defined healthcare context before broader implementation in diverse healthcare environments.

#### 2.2.1 Framework Overview for Operationalizing SDoH

Figure 2 provides a visual representation of our analytical framework for operationalizing SDoH in breast cancer screening programs. This framework, as detailed in Section 2.1, offered a structured approach to integrating diverse healthcare data sources, implementing appropriate machine learning models, validating predictive performance, generating explainable insights, and translating findings into future intervention strategies. Building upon the methodological foundation established by previous work on breast cancer screening prediction models (Sacca *et al*., 2024), we compared the performance of multiple machine learning techniques. These techniques provided analytical strengths while maintaining interpretability for healthcare practitioners.

**Figure 2:**
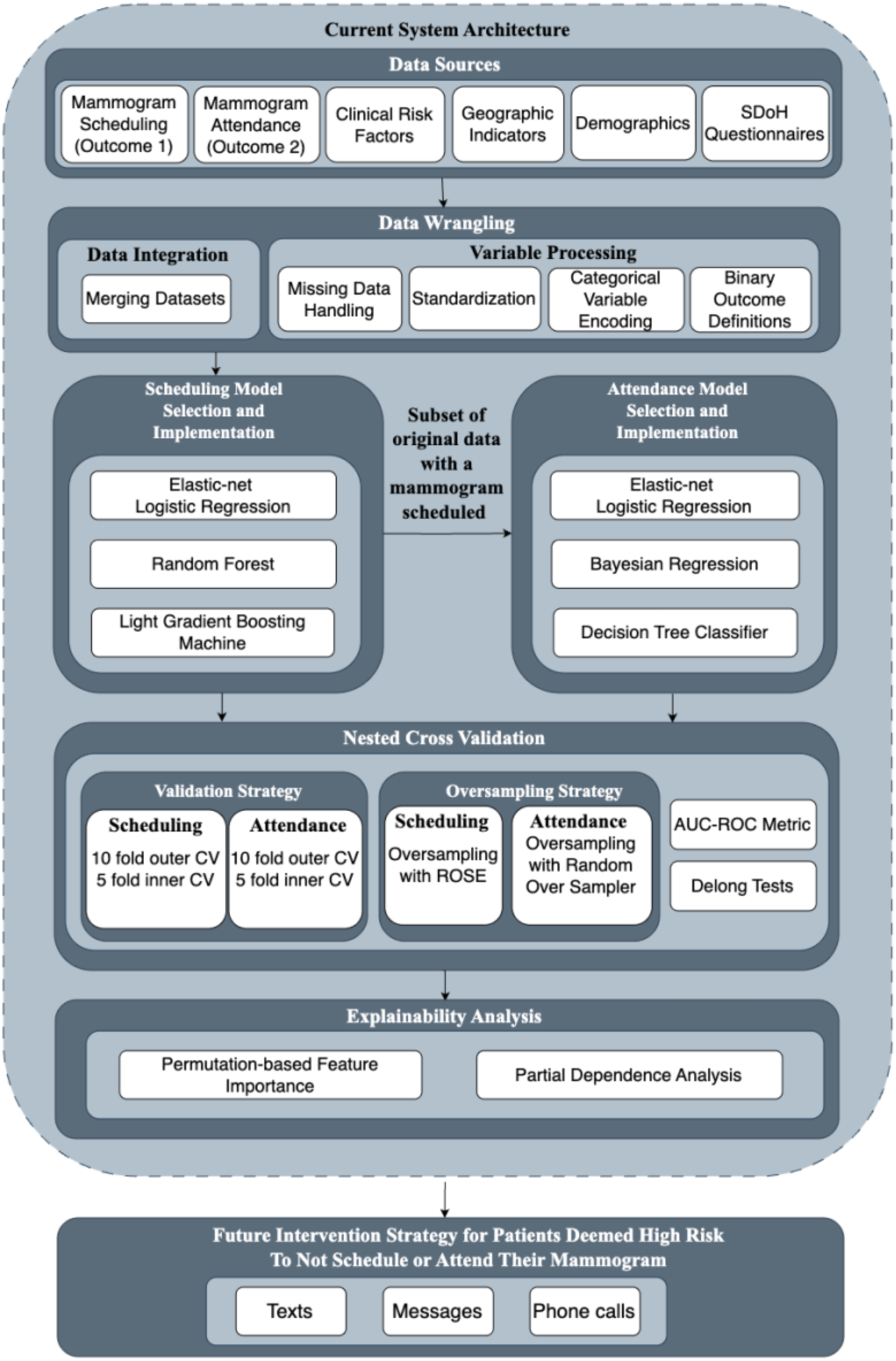
Framework Overview for Operationalizing SDoH in the Dartmouth Health System.

#### 2.2.2 Study Design and Data Sources

Our study built upon data from the Dartmouth Health Cancer Screening Outreach Program to develop a framework for operationalizing SDoH into breast cancer scheduling practice. This program operates within Dartmouth Health’s network, which spans primary and specialty care services throughout New Hampshire and Vermont. A notable characteristic of the Dartmouth Health dataset is its relatively limited racial and ethnic diversity, reflecting the demographic composition of northern New England. This relative homogeneity creates a more controlled environment for analyzing other social determinants affecting predominantly rural populations, though it may restrict the model’s ability to capture certain disparities related to race and ethnicity.

The study integrated three primary data sources: (1) patient information from individuals with established primary care relationships (n=63,537), defined as those who receive their usual primary care with Dartmouth Health and have had at least one visit with a Dartmouth Health primary care provider in the previous three years (medically-homed patients); (2) SDoH questionnaire responses (n=18,359) capturing various dimensions of patient health related social needs; and (3) clinical risk assessment scores from the Epic electronic health record systems, including the Charlson Comorbidity Index and general adult health scores, which provided standardized measures of patient health status and comorbidities. Our primary outcome measure was mammogram scheduling status and attendance status, which served as our indicators of patient engagement with breast cancer screening.

#### 2.2.3 Study Population and Inclusion Criteria

The study population encompassed female patients aged 50-75 years receiving active care at Dartmouth Health primary care clinics, with active care defined as having completed a primary care visit at the health system within the previous three years. To maintain focus on adherence to standard Dartmouth Health breast cancer screening schedules for women with average risk, we excluded patients with breast cancer history or elevated risk factors that would necessitate different scheduling protocols. To ensure consistent screening practices across study sites, we also excluded two clinical sites that utilized different appointment scheduling protocols from the standard Dartmouth Health approach. While these sites demonstrated higher adherence rates due to automatic scheduling, their inclusion would have confounded our analysis of standard care patterns by introducing scheduling protocol variability.

#### 2.2.4 Model Validation

To ensure the external validity of our findings, we employed our models on a hold-out test set (20% of the data) that was not used during model development or hyperparameter tuning. This approach provided an unbiased assessment of model generalizability to new patients within the Dartmouth Health System. We applied consistent performance evaluation metrics between our development and test phases, allowing us to directly compare predictive capabilities and quantify how effectively our models can identify screening patterns in previously unseen data. This evaluation on independent data helped determine whether the relationships identified during model training remained stable when applied in new contexts, providing healthcare systems with confidence that the implementation insights generated by our models would be reliable and actionable in clinical settings.

### 2.3 Sensitivity Analyses and Secondary Analyses

To assess the robustness of our findings to different analytical assumptions, we conducted sensitivity analyses focusing on missing data handling approaches. Specifically, we performed complete case analyses using only patients with complete SDoH questionnaire data as sensitivity checks for our primary imputation-based approach. These analyses used identical modeling frameworks and performance evaluation metrics as described in Section 2.1 to ensure comparability with our primary results.

Additionally, we conducted comprehensive secondary analyses to provide deeper insights into factors influencing mammogram screening behaviors, including age-stratified evaluations, SDoH-only models, patient-level models, and clinic-level analyses. Detailed methodologies and results for all secondary analyses are presented in Supplementary Materials A1.1-A1.5 (Scheduling analyses) and A2.1-A2.5 (Attendance analyses).

## 3. RESULTS

### 3.1 Data Structure and Missingness

Our analysis of SDoH questionnaire data revealed substantial variation in response completeness across the 37 administered questions. Missingness rates ranged from 10.2% to 92.4%, with a median missingness of 73.8% across all questions (Table A1). This evaluation identified 11 questions that exceeded our pre-established 80% missingness threshold, which were subsequently excluded from model development to ensure implementation reliability. The excluded questions primarily addressed sensitive domains such as mental health status, substance use behaviors, and detailed information regarding past scheduling experiences.

Examination of the dataset revealed distinct patterns in both scheduling and attendance behaviors. Scheduling rates, calculated as the proportion of all eligible women aged 50-75 who had a mammogram scheduled, showed substantial variation across clinical sites (4.4%-21.3%), insurance types (Medicare: 16%; Commercial: 12.9%), age groups, and neighborhood deprivation levels. For attendance, missed appointment rates varied by clinical site (1.9%-9.1%), insurance status (Medicaid Managed: 13%; Blue Cross: 4%), and racial demographics (Asian: 0.8%; Hispanic: 8.1%) (Table A4). We found a linear relationship between neighborhood deprivation and missed appointments (ADI 1: 1.6%; ADI 10: 11.4%) and higher attendance in urban areas compared to rural settings. SDoH questionnaire responses indicated that housing instability (multi-residence: 12.4% vs. single-residence: 4.8% missed appointments), transportation barriers (unable to work due to transportation: 18.8% vs. no barriers: 4.9%), food insecurity (often: 14.3% vs. never: 4.9%), and health literacy challenges were associated with lower scheduling rates and higher missed appointment rates (Tables A2, A3, A5).

### 3.2 Analytical Framework Performance

#### 3.2.1 Scheduling Model Performance

The light gradient boosting model demonstrated a moderate average out-of-sample performance in our cross-validation scheme (AUC=0.709), followed by random forest (AUC=0.702) and elastic-net logistic regression (AUC=0.608). Both tree-based models significantly outperformed the logistic regression approach, with the light gradient boosting model showing a statistical advantage over logistic regression (AUC difference=0.050, p<0.001) and random forest similarly demonstrating superior performance compared to logistic regression (AUC difference=0.05, p<0.001). The difference between gradient boosting and random forest models was minimal (AUC difference=0.0001) and not statistically significant (p=0.972), confirming that both tree-based approaches had comparable predictive power for this scheduling behavior prediction.

The gradient boosting model, our best-performing approach, showed strong consistency across validation scenarios. The model’s performance ranged from 0.707 (worst AUC on validation sets) to 0.711 (best AUC on validation sets), indicating stable predictive performance. Our AUC on the held-out test set, which predicts model performance on unseen data, also achieved a relatively similar AUC of 0.67. This stability was particularly important for healthcare systems implementing SDOH-informed scheduling programs across diverse communities.

#### 3.2.2 Attendance Model Performance

For attendance prediction, we used an identical approach to compare three machine learning models: Bayesian regression (AUC=0.702), elastic-net logistic regression (AUC=0.699), and decision tree classifier (AUC=0.666).

Delong’s test showed that these three models performed comparably (AUC difference Bayes-Log: 0.004, p > 0.05) (AUC difference bayes-tree: 0.00423, p >0.05) (AUC difference log-tree: 0.00323, p >0.05). Given this comparable performance, we selected logistic regression as our final model for its computational simplicity and independence from prior assumptions. This selected model demonstrated moderate consistency across validation datasets. Performance ranged from AUC = 0.6531 to AUC = 0.7851, with an average validation AUC of 0.7282. When evaluated on the held-out test set, the model maintained robust performance (AUC = 0.699), which showed somewhat consistent predictive power.

#### 3.2.3 Permutation-Based Variable Importance

Our variable importance analysis from light gradient boosting machine (scheduling model) and elastic-net logistic regression (attendance model) using a permutation-based approach identified key social drivers for future implementation focus (Figure 3).

**Figure 3:**
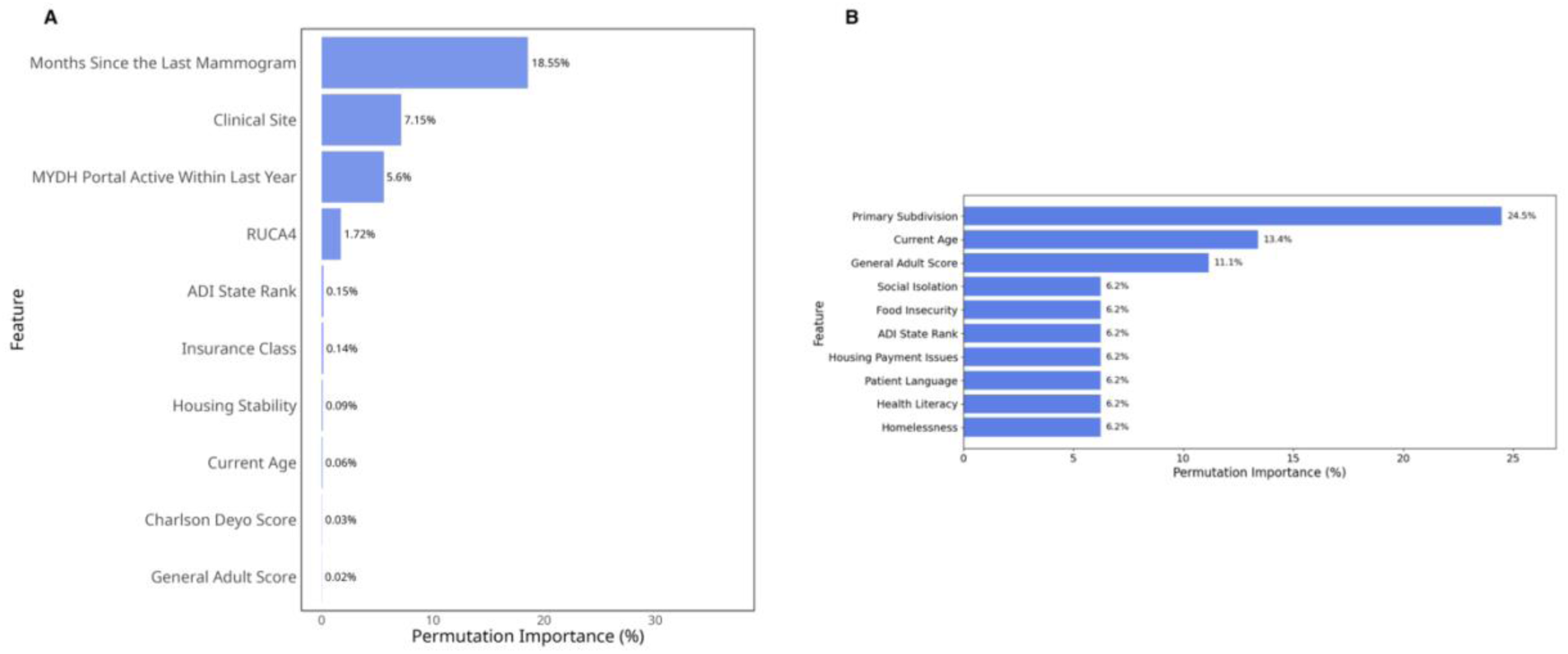
Permutation Importance based on Percentage Decrease in AUC. (A) Top 10 most important variables in the scheduling model; (B) Top 10 most important variables in the attendance model.

For the scheduling model (Figure 3A), months since the last mammogram emerged as the strongest individual predictor with a permutation importance value of 18.55%. Clinical site was the second most influential factor (7.15%), followed by MYDH Portal active within last year (5.6%). When considering cumulative effects, these top three features together represented more than 30% of the total permutation importance, suggesting that temporal, demographic, and organizational factors were particularly crucial for non-scheduling behavior. Geographic and socioeconomic factors also showed some influence, with RUCA4 (1.72%) and ADI state rank (0.15%) completing the top five predictors. Almost all traditional SDoH questionnaire responses such as homelessness (0.01%), food insecurity (0.01%), and financial hardship (0.01%) showed limited predictive power in our model and were therefore excluded from the diagram. This less prominent role of direct SDoH questionnaire measures compared to geographic and facility-level indicators suggested that social determinants may exert their influence through complex pathways that are better captured by community-level metrics and healthcare delivery characteristics than by individual self-reported social needs.

For the attendance model, clinical site is the most influential variable, contributing 24.5% to model performance, followed by current age (13.4%) and general adult score (11.1%). This importance indicates that both site-level factors and patient health burden strongly influence attendance (Figure 3B). Social Isolation (6.2%), Food Insecurity (6.2%), and ADI State Ranks (7.4%) also played notable roles, suggesting that social interaction, resource availability, and geographic context needs affect screening adherence. In contrast to the scheduling model, the attendance model excluded months since the last mammogram (the strongest scheduling predictor) to avoid data leakage, as temporal information was incorporated into the attendance outcome definition (see Methods 2.1.1).

#### 3.2.4 Partial Dependence Plots

To further examine how key social drivers influence breast cancer screening behavior, we plotted partial dependence plots for the most influential predictors (Figure 4). For the scheduling model, months since the last mammogram showed a distinct temporal pattern with particularly higher probability of not scheduling within the first few months, followed by a significant drop around 10-12 months, and subsequent fluctuations that stabilize after approximately 30 months (Figure 4A). Among categorical predictors, clinical site demonstrated some variation in the probability of not scheduling across different healthcare facilities, with relatively consistent predicted probabilities ranging between approximately 0.45 and 0.55 (Figure 4B).

**Figure 4:**
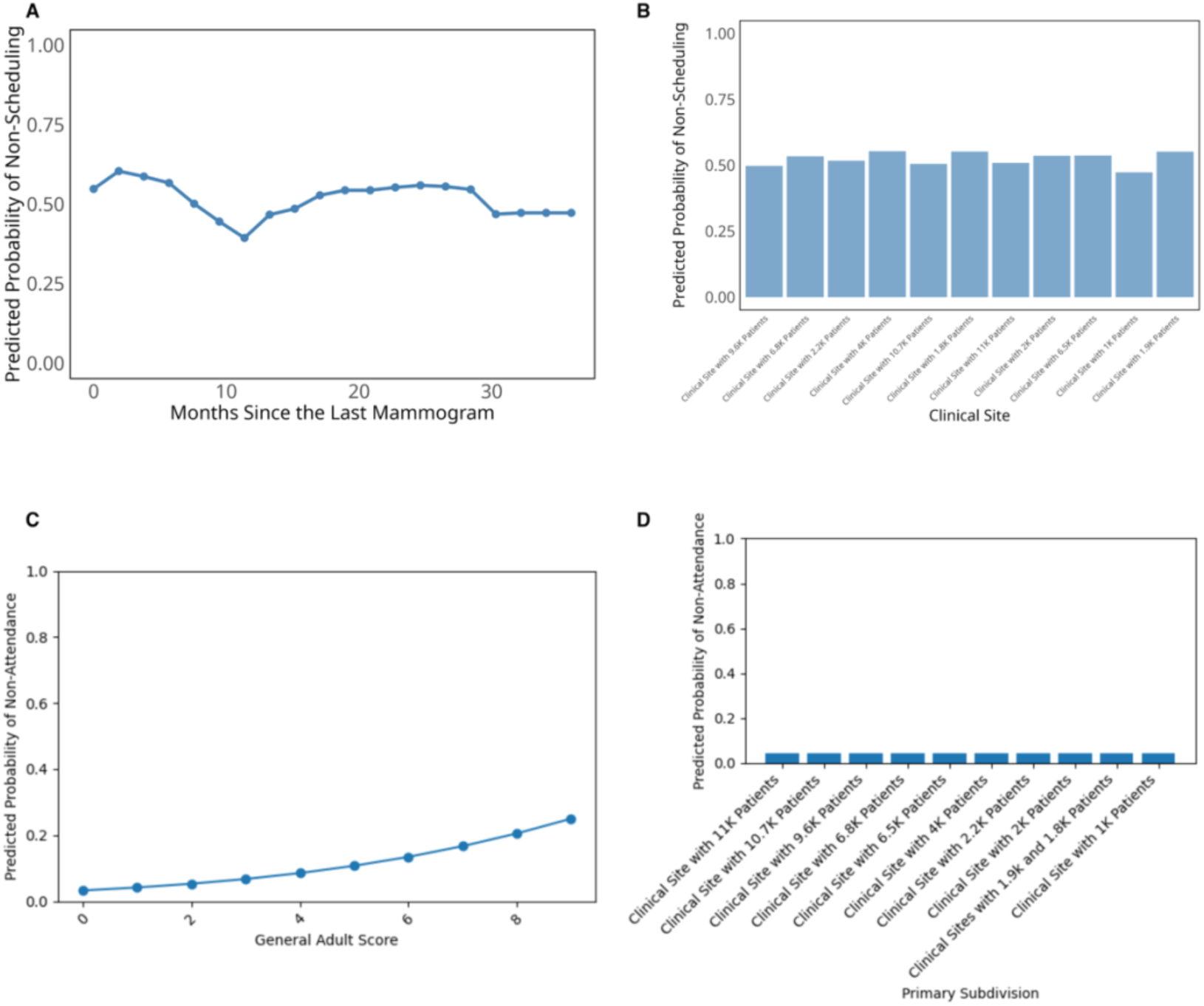
Partial Dependence Plots for Top Predictors in Breast Cancer Scheduling and Attendance Prediction. (A) Top numerical variable in the scheduling model; (B) Top categorical variable in the scheduling model; (C) Top numerical variable in the attendance model; (D) Top categorical variable in the attendance model.

For the attendance model (Figure 4C-D), the general adult score showed a clear positive relationship with non-attendance probability, with higher scores indicating greater health complexity and comorbidity burden. These higher scores were associated with increased likelihood of missing scheduled appointments, rising from near-zero probability at low scores to approximately 0.25 at the highest health complexity scores (Figure 4C). In contrast, clinical site showed minimal variation in attendance patterns, with predicted non-attendance probabilities remaining consistently low (below 0.1) across most healthcare facilities (Figure 4D). While clinical site was one of the top important variables in our variable importance analysis, the practical differences in attendance rates between sites were modest once patient-level factors are accounted for.

Additional variables examined in our analysis, including Charlson Comorbidity Index, housing stability, patient race, age, and various social determinants of health measures showed relatively minimal impact on mammography screening or little variation in not scheduling (Figure A1) and screening non-attendance probability (Figures A2).

## 4. DISCUSSION

Our study showed that machine learning approaches can effectively identify the factors that influence breast cancer scheduling and attendance behavior within a healthcare system. While we initially examined SDoH as potential drivers of screening patterns, our findings revealed that healthcare systems may achieve better impact by focusing on factors within their direct control. The light gradient boosting model achieved clinically meaningful performance comparable to other predictive models addressing SDoH-related scheduling outcomes (Sotudian *et al*., 2022; Stabellini *et al*., 2023). Furthermore, the elastic-net logistic regression model achieved modest performance relative to other predictive models when addressing SDoH related attendance outcomes (Nelson *et al*., 2019; Salazar *et al*., 2020). Our findings highlighted several key implementation domains: temporal patterns in scheduling behavior revealed the dynamic nature of patient engagement; facility-level variables emerged as important predictors, reflecting the influence of organizational characteristics; and social determinants and geographic factors demonstrated the impact of community context, though to a lesser degree than anticipated. While the model’s performance reflected the inherent challenges of quantifying social factors, it may provide healthcare systems with actionable insights for implementing scheduling programs that address both organizational and community-level barriers while accounting for individual patient characteristics.

### 4.1 Implementation Implications

Our analysis revealed that the relationship between social determinants and screening behavior involved multiple interacting factors. The light gradient boosting model’s ability to capture these relationships (AUC=0.709), together with similar performance from the random forest approach (AUC=0.702), suggested that accounting for non-linear interactions between social drivers may help healthcare systems better understand scheduling behavior patterns.

Consistent with established literature, our models confirmed that geographic accessibility (RUCA) and socioeconomic factors (ADI state rank) influence screening behaviors (Miller *et al*., 2019; Pohl *et al*., 2025). However, our variable importance analysis revealed a hierarchy of influence that differs from traditional approaches focused primarily on individual-level social barriers. Temporal factors (months since last mammogram) and organizational factors (clinical site) emerged as the strongest predictors, suggesting that healthcare systems may achieve more immediate impact through system-level interventions rather than attempting to address individual patients’ social circumstances. This finding highlights a different priority than much of the existing mammography literature, which emphasizes individual-level barriers such as transportation, cultural beliefs, and health literacy (Lofters *et al*., 2017; Ponce-Chazarri *et al*., 2023; Albadawi, Alsharawneh and Othman, 2025). While these individual SDoH factors remained important in our descriptive analyses, our machine learning approach revealed that facility-level variations and care patterns were more predictive of scheduling and attendance behavior. For Dartmouth Health specifically, this suggests that standardizing practices across clinical sites may yield greater improvements in screening rates than traditional patient-education or transportation-assistance programs. For attendance behavior, the evidence suggests a dual approach combining system-level standardization with targeted interventions. This recommendation is supported by our clinic-level analysis for scheduling (A1.4), which confirmed substantial performance variation across sites. However, the clinic-level analysis for attendance (A2.4) showed inconsistent results, limiting conclusions about organizational effects.

Our findings also illuminate the complex interplay between organizational and social factors that traditional regression approaches often miss (Coughlin, 2019). The elastic-net logistic regression model’s performance in capturing attendance patterns (AUC=0.698) demonstrated that patient health complexity (general adult score) and digital engagement (portal activity) are critical factors that complement traditional socioeconomic predictors. This insight provides healthcare systems with a more nuanced understanding of how to target interventions across different patient populations.

### 4.2 Methodological Contributions

Our analytical approach offered several methodological contributions to healthcare delivery and the implementation science. First, we demonstrated a novel approach to operationalizing SDoH in breast cancer scheduling and attendance practices, providing healthcare systems with a framework to translate social determinant screening tools into actionable screening strategies. Unlike previous work analyzing nationwide census tract-level scheduling rates and focused primarily on geographic accessibility and demographics, our study examined individual-level data integrating clinical, behavioral, and social determinants within a healthcare system context (Hashtarkhani *et al*., 2025). This focus allowed us to identify specific patient-level factors that directly influence scheduling decisions, rather than ecological correlations at the population level.

Second, our modeling framework effectively functioned as a poly-social risk score system, aggregating multiple social determinants to quantify their combined influence on screening adherence. This approach moved beyond examining isolated social determinants to consider how they collectively impact health behaviors. Third, the age-stratified analysis (Supplementary Material A1.2) revealed important variations in these poly-social risk profiles across demographic groups, suggesting the need for age-specific implementation strategies that account for different SDoH impacts across the lifespan. Finally, our application of light gradient boosting models and elastic-net logistic regression models and partial dependence plots revealed important non-linear patterns in the relationship between months since the last mammogram and non-scheduling probability—a critical insight that traditional regression approaches would likely miss. For the attendance model, our elastic-net logistic regression approach similarly captured complex relationships between organizational factors, patient characteristics, and social determinants, though with different key predictors than the scheduling model. These findings demonstrate the value of machine learning approaches in capturing complex relationships between certain social determinants and screening behavior.

The development of a unified modeling framework that incorporated both individual-level social drivers and system-level factors provided healthcare organizations with a template for analyzing their own screening programs. This approach could be particularly valuable as healthcare systems work to improve cancer screening rates for their medically-homed populations while effectively integrating SDoH data into their quality improvement initiatives.

### 4.3 Future Directions in Implementation Science

While our study provides valuable quantitative insights into the relationship between social determinants and breast cancer screening behaviors, future work should consider how these findings could inform comprehensive implementation science approaches. The Consolidated Framework for Implementation Research (CFIR) offers a valuable lens for future efforts to translate our findings into practice. Though our current work focuses on quantitative modeling rather than a full CFIR implementation, our findings provide a foundation for subsequent mixed-methods approaches that could more fully leverage implementation science frameworks.

For example, the facility-level variations identified in our model align with CFIR’s ‘inner setting’ domain, suggesting that organizational culture and readiness for implementation play important roles in both scheduling and attendance behaviors. Future work could build on our quantitative findings by using qualitative methods to explore how these organizational factors influence practices and how interventions might be tailored to different clinical settings. Similarly, our findings related to geographic and socio-economic factors correspond to CFIR’s ‘outer setting’ domain, highlighting the importance of understanding patient needs and resources with their community investigations could provide deeper insights into how these community factors shape decisions and how healthcare systems might better address them.

As healthcare systems consider implementing SDoH-informed interventions, CFIR and other implementation science frameworks could provide valuable guidance for assessing feasibility, sustainability, and potential barriers. Our work represents an important first step in this direction by providing quantitative evidence of key relationships that future implementation efforts should consider.

### 4.4 Future Research Priorities

Our model was developed and validated within the Dartmouth Health system, which serves a population with limited racial, ethnic, and linguistic diversity. This demographic homogeneity may have limited our ability to capture important language-related barriers to screening access and communication, and may have restricted the generalizability of our findings to more diverse healthcare settings and populations. Although our findings indicate small racial differences, future work should validate these approaches in healthcare systems serving more diverse communities to ensure broader applicability. It is important to note, however, that we have designed our work as a generalizable framework that can perform well in other situations and can incorporate more diverse racial groups and other demographic aspects if the necessary data are available.

Moreover, our modeling approach assumed that the ratio between screening and non-screening populations will remain stable over time. This assumption may not hold in different implementation contexts or as scheduling programs evolve. Healthcare systems implementing similar approaches should carefully consider their local population characteristics and mammogram scheduling patterns. Additionally, while our model demonstrated modest predictive performance within our system, its generalizability to other healthcare settings may be limited by differences in organizational structure, population characteristics, screening protocols, and the substantial missingness in our SDoH data, which may have limited our ability to fully capture social determinant influences. Future research should explore how these models can be adapted and calibrated for different healthcare contexts.

Beyond expanding the population and removing assumptions, we identified several priority areas for future research. First, the development of dynamic modeling approaches that can adapt to changing population characteristics and scheduling patterns would enhance the robustness of our framework. Additionally, integrating SDoH-informed scheduling models with other preventive care programs could create more comprehensive implementation strategies. Investigation of facility-level variations in scheduling patterns would have further identified best practices for implementation. Finally, extending our analytical framework to other scheduling programs, such as colorectal and cervical cancer scheduling, would increase the broader applicability of SDoH-informed modeling approaches and strengthen the overall impact of our generalizable framework across diverse healthcare settings.

### 4.5 Conclusion

Our study provided healthcare systems with a data-driven approach to understanding and addressing how social determinants shape breast cancer scheduling practices. Our findings suggested that machine learning approaches could help healthcare systems develop more effective, targeted implementation strategies. As healthcare systems work to meet cancer screening targets for their medically-homed populations, approaches that systematically analyze and address social determinants of health could become increasingly valuable for improving adherence and reducing disparities.

Looking ahead, our quantitative findings provide a foundation for future implementation science approaches that could more fully leverage frameworks like CFIR to translate these insights into practice. By combining machine learning approaches with implementation science, healthcare systems can develop more comprehensive strategies for addressing the complex interplay between social determinants and screening behaviors, ultimately improving health outcomes for diverse patient populations.

## Data Availability

All data produced in the present study are available upon reasonable request to the authors

## 5. DECLARATION

### 5.1 Ethics statement

This study is classified as exempt by the Dartmouth Health Institutional Review Board. All data collection and analysis procedures comply with relevant privacy regulations and institutional policies for handling protected health information. The requirement for individual patient consent was waived due to the use of de-identified data.

### 5.3 Author disclaimer

The contents of this work are the authors’ sole responsibility and do not necessarily represent the official views of Dartmouth Health.

## SUPPLEMENTARY FILES

### Supplementary Figures

**Figure A1:**
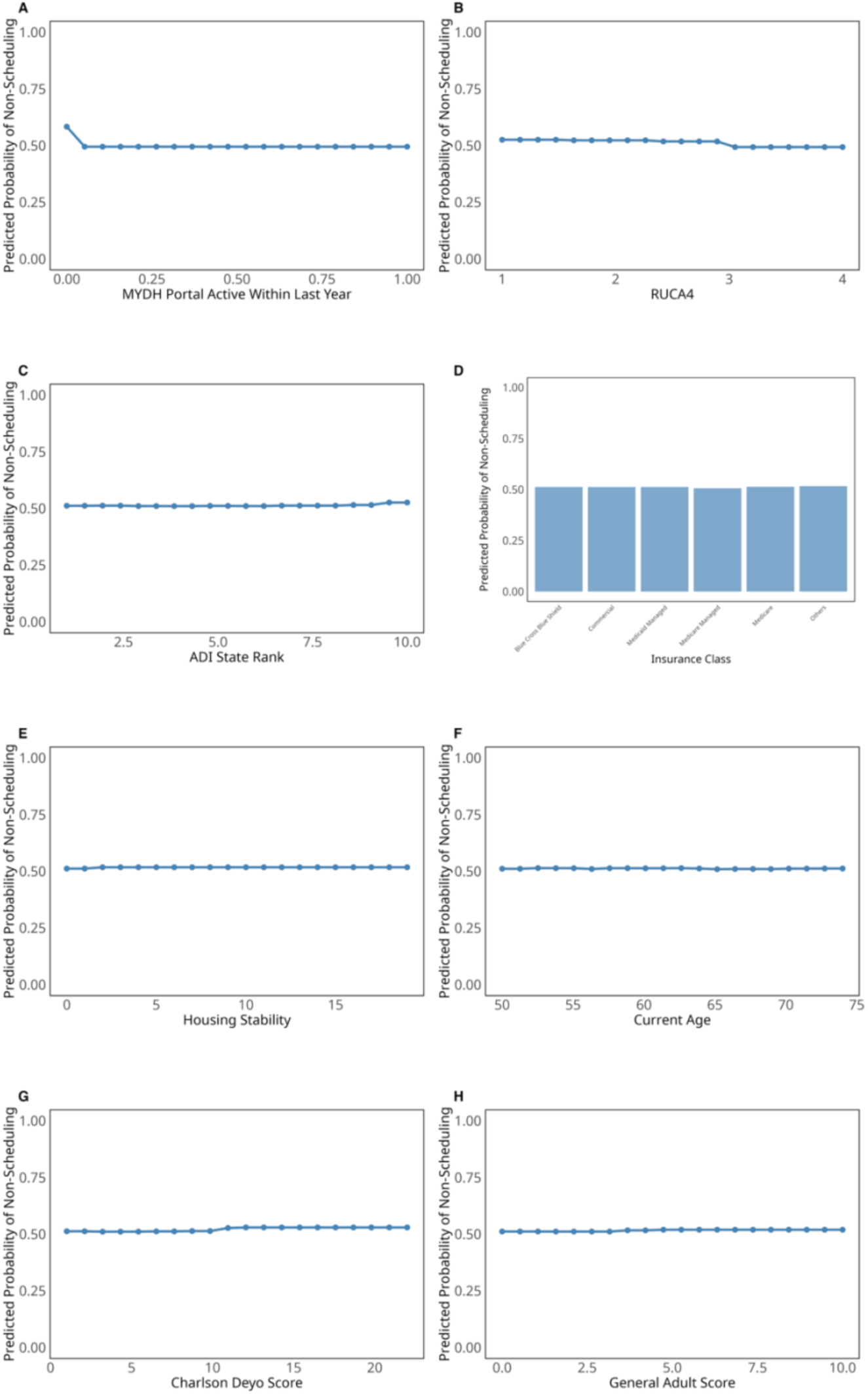
Partial Dependence Plots for Additional Variables. The plots display the relationship between various factors and mammography scheduling probability: (A) MYDH Portal Active Within Last Year, (B) RUCA-4, (C) ADI State Rank; (D) Insurance Class; (E) Housing Stability; (F) Current Age; (G) Charlson Deyo Score; (H) General Adult Score. Most of these variables show minimal influence on scheduling probability, with relatively flat relationships compared to the stronger predictors shown in Figure 3 and 4.

**Figure A2:**
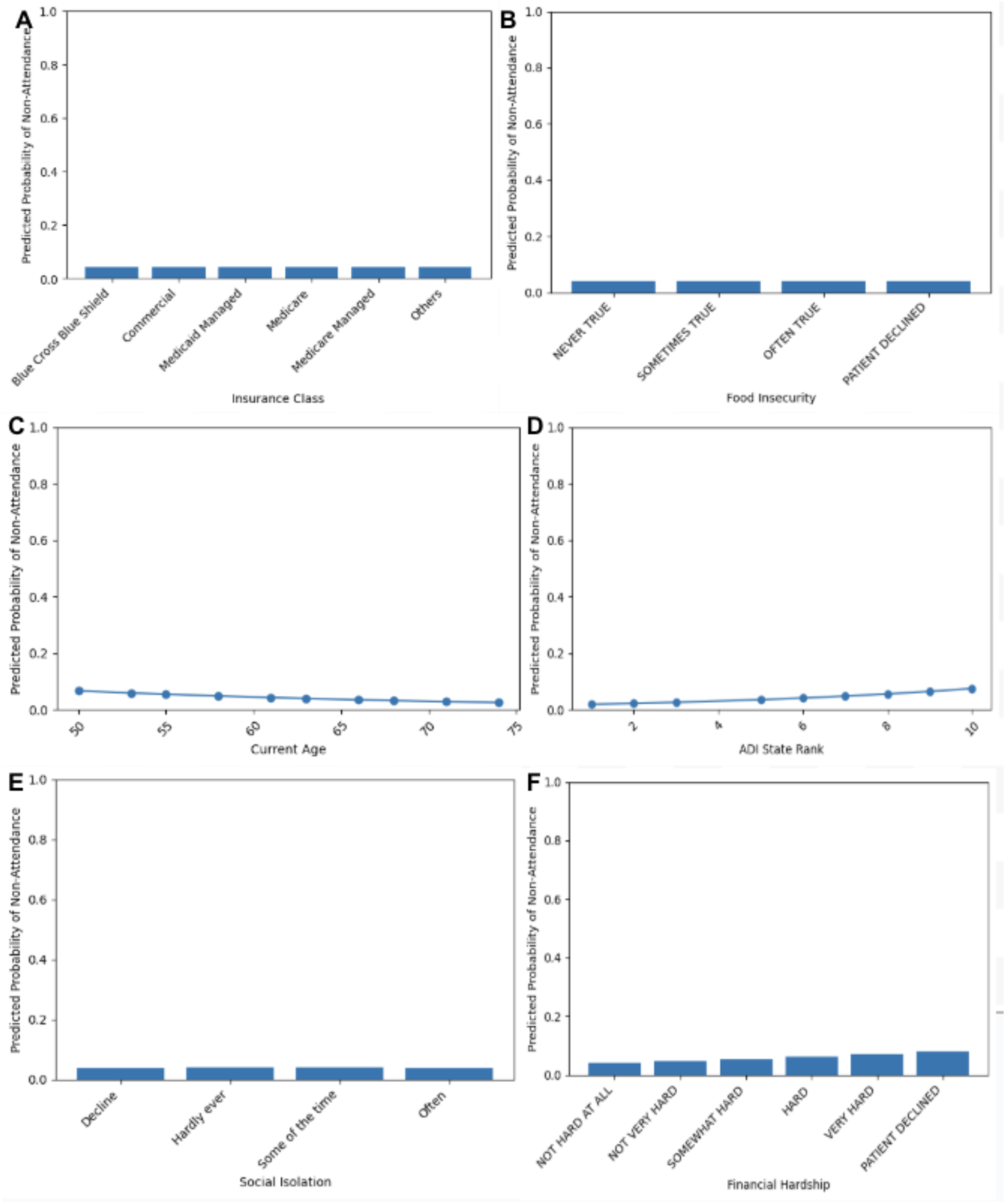
Partial Dependence Plots for Additional Variables. The plots display the relationship between various factors and mammography scheduling probability: (A) MYDH Portal Active Within Last Year, (B) RUCA-4, (C) Current Age, (D) Patient Race, (E)Insurance Class, (F) Financial Hardship.

### Supplementary Tables

**Table A1:** Missingness Per Question. Missing Percentage Per Question (%) is calculated as (Missing Responses/Total Eligible Patients)*100. Missing responses include both patients who never encounter the question and those who encountered it but did not provide a valid response (excluding ‘Patient Refused’ and ‘Declined’ which were retained as meaningful responses.

| QUESTION | Number of Responses | Total Responses | Evaluable Responses | Missing Percentage Per Question (%) |
| --- | --- | --- | --- | --- |
| How hard is it for you to pay for the very basics like food, housing, medical care, and heating? | 18357 | 18583 | 226 | 1.22 |
| Within the past 12 months, you worried that your food would run out before you got the money to buy more. | 18313 | 18548 | 235 | 1.27 |
| In the last 12 months, was there a time when you were not able to pay the mortgage or rent on time? | 18277 | 18568 | 291 | 1.57 |
| In the past 12 months, has lack of transportation kept you from meetings, work, or from getting things needed for daily living? | 18145 | 18709 | 564 | 3.01 |
| In the past 12 months, has lack of transportation kept you from medical appointments or from getting medications? | 18174 | 18801 | 627 | 3.33 |
| How often do you feel isolated from others? | 18183 | 18883 | 700 | 3.71 |
| In the last 12 months, was there a time when you did not have a steady place to sleep or slept in a shelter (including now)? | 17761 | 18633 | 872 | 4.68 |
| Within the past 12 months, the food you bought just didn't last and you didn't have money to get more. | 18295 | 19216 | 921 | 4.79 |
| How often do you need to have someone help you when you read instructions, pamphlets, or other written material from your doctor or pharmacy? | 17681 | 18667 | 986 | 5.28 |
| In the last 12 months, how many places have you lived? | 17787 | 19819 | 2032 | 10.25 |
| In the past 12 months has the electric, gas, oil, or water company threatened to shut off services in your home? | 9575 | 18517 | 8942 | 48.29 |
| How often do you feel that you lack companionship? | 3528 | 18486 | 14958 | 80.92 |
| How often do you feel left out? | 3524 | 18476 | 14952 | 80.93 |
| What was your main activity during most of the last 12 months? | 3509 | 18442 | 14933 | 80.97 |
| Do you have any legal issues that you are having difficulty resolving, such as disability, custody, parole, eviction, restraining orders, or other? | 3507 | 18450 | 14943 | 80.99 |
| Do you feel unsafe in your relationships at home, school or work? | 3502 | 18444 | 14942 | 81.01 |
| Has someone threatened to hurt you, your family or your pets? | 3499 | 18449 | 14950 | 81.03 |
| Have you ever felt that someone wants to hurt you? | 3495 | 18438 | 14943 | 81.04 |
| Does anyone try to keep you from having contact with others or doing things outside your home? | 3495 | 18441 | 14946 | 81.05 |
| Do you have reliable use of a phone? | 3483 | 18424 | 14941 | 81.1 |
| Do you have reliable access to the internet? | 3480 | 18427 | 14947 | 81.11 |
| Social Isolation Score | 3484 | 18645 | 15161 | 81.31 |
| Are you able to speak on the phone in a safe and private manner? | 3403 | 18446 | 15043 | 81.55 |
| Are you able to access the internet in a safe and private manner? | 3336 | 18444 | 15108 | 81.91 |
| Within the last year, have you been humiliated or emotionally abused in other ways by your partner or ex-partner? | 1878 | 18377 | 16499 | 89.78 |
| Within the last year, have you been afraid of your partner or ex-partner? | 1874 | 18379 | 16505 | 89.8 |
| Within the last year, have you been raped or forced to have any kind of sexual activity by your partner or ex-partner? | 1872 | 18379 | 16507 | 89.81 |
| Within the last year, have you been kicked, hit, slapped, or otherwise physically hurt by your partner or ex-partner? | 1870 | 18379 | 16509 | 89.83 |
| What do you have trouble paying for? | 654 | 18711 | 18057 | 96.5 |
| At any time in the past 12 months, were you homeless or living in a shelter (including now)? | 476 | 18364 | 17888 | 97.41 |
| How often do you need to have someone help you when you read instructions, pamphlets, or other written material from your doctor or pharmacy? | 474 | 18365 | 17891 | 97.42 |
| In the past 12 months, how many times have you moved where you were living? | 476 | 18470 | 17994 | 97.42 |
| How hard is it for you to pay for the very basics like food, housing, medical care, and heating? | 1 | 18359 | 18358 | 99.99 |
| In the past 12 months, has lack of transportation kept you from medical appointments or from getting medications? | 1 | 18359 | 18358 | 99.99 |
| In the past 12 months, has lack of transportation kept you from meetings, work, or getting things needed for daily living? | 1 | 18359 | 18358 | 99.99 |
| Within the past 12 months, the food you bought just didn't last and you didn't have money to get more. | 1 | 18359 | 18358 | 99.99 |
| Within the past 12 months, you worried that your food would run out before you got money to buy more. | 1 | 18359 | 18358 | 99.99 |

**Table A2:** Consolidated Demographic Counts.

| Category | Number Not Scheduled | Number Scheduled | Proportion Not Scheduled |
| --- | --- | --- | --- |
| Clinical Site |  |  |  |
| Clinical Site with 11K Patients | 1878 | 304 | 86% |
| Clinical Site with 10.7K Patients | 537 | 130 | 80% |
| Clinical Site with 9.6K Patients | 804 | 212 | 79% |
| Clinical Site with 6.8K Patients | 855 | 109 | 89% |
| Clinical Site with 6.5K Patients | 964 | 145 | 87% |
| Clinical Site with 4K Patients | 240 | 21 | 92% |
| Clinical Site with 2.2K Patients | 192 | 38 | 83% |
| Clinical Site with 2K Patients | 273 | 34 | 89% |
| Clinical Site with 1.9K Patients | 306 | 14 | 96% |
| Clinical Site with 1.8K Patients | 394 | 27 | 94% |
| Clinical Site with 1K Patients | 100 | 27 | 79% |
| **Insurance Class** |  |  |  |
| Blue Cross Blue Shield | 2517 | 396 | 86% |
| Commercial | 1742 | 257 | 87% |
| Medicare | 949 | 181 | 84% |
| Medicare Managed | 866 | 158 | 85% |
| Others | 316 | 40 | 89% |
| Medicaid Managed | 153 | 29 | 84% |
| **Race** |  |  |  |
| White | 6021 | 979 | 86% |
| Others | 224 | 32 | 87% |
| Asian | 136 | 25 | 84% |
| Hispanic | 108 | 19 | 85% |
| Black | 54 | 6 | 90% |
| **Language** |  |  |  |
| English | 6490 | 1050 | 86% |
| Others | 53 | 11 | 83% |
| **Charlson Deyo Score** |  |  |  |
| 1 | 184 | 33 | 85% |
| 2 | 1767 | 264 | 87% |
| 3 | 2028 | 297 | 87% |
| 4 | 1121 | 197 | 85% |
| 5 | 622 | 139 | 82% |
| 6 | 342 | 53 | 87% |
| 7 | 159 | 24 | 87% |
| 8 | 100 | 24 | 81% |
| 9 | 67 | 11 | 86% |
| 10 | 46 | 6 | 88% |
| 11 | 37 | 4 | 90% |
| 12 | 36 | 6 | 85% |
| 13 | 11 | 1 | 91% |
| 14 | 12 | 1 | 92% |
| 15 | 4 | 0 | 100% |
| 16 | 4 | 0 | 100% |
| 17 | 1 | 0 | 100% |
| 18 | 1 | 0 | 100% |
| 19 | 1 | 1 | 40% |
| **General Adult Score** |  |  |  |
| 0 | 2540 | 350 | 88% |
| 1 | 2539 | 435 | 85% |
| 2 | 963 | 189 | 84% |
| 3 | 336 | 65 | 84% |
| 4 | 110 | 18 | 86% |
| 5 | 30 | 4 | 88% |
| 6 | 14 | 0 | 100% |
| 7 | 6 | 0 | 100% |
| 8 | 3 | 0 | 100% |
| 9 | 1 | 0 | 100% |
| 10 | 1 | 0 | 100% |
| **MyDH Active Within Last Year** |  |  |  |
| Yes | 6392 | 1050 | 86% |
| No | 151 | 11 | 93% |
| **Age Group** |  |  |  |
| 50-54 | 1388 | 216 | 87% |
| 55-59 | 1480 | 232 | 86% |
| 60-64 | 1601 | 223 | 88% |
| 65-69 | 1269 | 229 | 85% |
| 70-74 | 805 | 161 | 83% |
| **State Rank (ADI)** |  |  |  |
| 1 | 808 | 118 | 87% |
| 2 | 763 | 115 | 87% |
| 3 | 788 | 121 | 87% |
| 4 | 737 | 101 | 88% |
| 5 | 725 | 117 | 86% |
| 6 | 671 | 116 | 85% |
| 7 | 578 | 122 | 83% |
| 9 | 516 | 96 | 84% |
| 8 | 576 | 84 | 87% |
| 10 | 381 | 71 | 84% |
| **Rural-Urban Commuting Area** |  |  |  |
| 1 | 2408 | 359 | 87% |
| 2 | 443 | 62 | 88% |
| 3 | 2355 | 396 | 86% |
| 4 | 1337 | 244 | 85% |

**Table A3:** Consolidated Question Counts.

| Category | Number Scheduled | Proportion Scheduled | Number Not Scheduled | Proportion Not Scheduled |
| --- | --- | --- | --- | --- |
| **Unable to pay rent or mortgage in the past 12 months** |  |  |  |  |
| No | 987 | 0.1407 | 6026 | 0.8592 |
| Yes | 42 | 0.1183 | 313 | 0.8814 |
| Patient Declined | 16 | 0.2133 | 59 | 0.7844 |
| Patient Refused | 16 | 0.0994 | 145 | 0.9001 |
| **Number of residences in the past 12 months** |  |  |  |  |
| 0 | 8 | 0.1905 | 34 | 0.8059 |
| 1 | 986 | 0.1397 | 6074 | 0.8603 |
| 2 | 59 | 0.1379 | 369 | 0.8619 |
| 3 | 6 | 0.1132 | 47 | 0.8849 |
| 4 | 0 | 0.0 | 8 | 1.0 |
| 5 | 0 | 0.0 | 2 | 1.0 |
| 7 | 0 | 0.0 | 2 | 1.0 |
| 9 | 0 | 0.0 | 1 | 1.0 |
| 10 | 0 | 0.0 | 2 | 1.0 |
| 12 | 1 | 0.5 | 1 | 0.4 |
| 13 | 0 | 0.0 | 2 | 1.0 |
| 19 | 1 | 1.0 | 0 | 0.0 |
| 33 | 0 | 0.0 | 1 | 1.0 |
| **Unable to access medication due to transportation difficulties** |  |  |  |  |
| No | 1033 | 0.1397 | 6363 | 0.8603 |
| Yes | 18 | 0.1731 | 86 | 0.8255 |
| Patient Declined | 10 | 0.0962 | 94 | 0.903 |
| **Feels isolated from others** |  |  |  |  |
| Hardly ever | 863 | 0.1419 | 5219 | 0.8581 |
| Some of the time | 133 | 0.1329 | 868 | 0.867 |
| Often | 35 | 0.1311 | 232 | 0.8685 |
| Decline | 30 | 0.1181 | 224 | 0.8815 |
| **Difficulty paying for necessities** |  |  |  |  |
| Not Hard at All | 721 | 0.1424 | 4343 | 0.8576 |
| Not Very Hard | 197 | 0.1315 | 1301 | 0.8684 |
| Somewhat Hard | 93 | 0.1418 | 563 | 0.858 |
| Patient Declined | 20 | 0.1149 | 154 | 0.8845 |
| Hard | 16 | 0.1345 | 103 | 0.8646 |
| Very Hard | 14 | 0.1505 | 79 | 0.8481 |
| **Concerns about inadequate access to food in the past 12 months** |  |  |  |  |
| Never True | 984 | 0.1411 | 5990 | 0.8589 |
| Sometimes True | 44 | 0.1239 | 311 | 0.8758 |
| Patient Declined | 20 | 0.1036 | 173 | 0.8959 |
| Often True | 13 | 0.1585 | 69 | 0.8398 |
| **Difficult reading medical material** |  |  |  |  |
| Never | 965 | 0.1386 | 5997 | 0.8614 |
| Rarely | 49 | 0.1546 | 268 | 0.845 |
| Sometimes | 21 | 0.1338 | 136 | 0.8655 |
| Decline | 11 | 0.1358 | 70 | 0.8628 |
| Always | 8 | 0.1905 | 34 | 0.8059 |
| Often | 7 | 0.1556 | 38 | 0.8415 |
| **Lacked a consistent residence in the past 12 months** |  |  |  |  |
| No | 1044 | 0.1405 | 6389 | 0.8595 |
| Yes | 8 | 0.1026 | 70 | 0.8963 |
| Patient Refused | 5 | 0.0769 | 60 | 0.922 |
| Patient Declined | 4 | 0.1429 | 24 | 0.8528 |
| **Unable to work due to lack of transportation** |  |  |  |  |
| No | 1036 | 0.1401 | 6357 | 0.8599 |
| Yes | 15 | 0.1562 | 81 | 0.8424 |
| Patient Declined | 10 | 0.087 | 105 | 0.9124 |
| **Insufficient access to food in the past 12 months** |  |  |  |  |
| Never True | 991 | 0.1399 | 6091 | 0.8601 |
| Sometimes True | 40 | 0.1515 | 224 | 0.848 |
| Patient Declined | 21 | 0.1082 | 173 | 0.8913 |
| Often True | 9 | 0.1406 | 55 | 0.8575 |

**Table A4:** Consolidated Demographic Counts-- Attendance.

| Category | Number Not Attended | Number Attended | Proportion Not Attended |
| --- | --- | --- | --- |
| **Clinical Site** |  |  |  |
| Clinical Site with 11K Patients | 62 | 1192 | 4.94% |
| Clinical Site with 10.7K Patients | 117 | 1176 | 9.05% |
| Clinical Site with 9.6K Patients | 10 | 1202 | 0.83% |
| Clinical site with 6.8K Patients | 19 | 446 | 4.09% |
| Clinical Site with 6.5K Patients | 26 | 549 | 4.52% |
| Clinical Site with 4K Patients | 16 | 337 | 4.53% |
| Clinical Site with 2.2K Patients | 19 | 256 | 6.91% |
| Clinical Site with 2K Patients | 8 | 125 | 6.02% |
| Clinical Site with 1.9K Patients | 5 | 82 | 5.75% |
| Clinical Site with 1.8K Patients | 6 | 91 | 6.19% |
| Clinical Site with 1K Patients | 2 | 104 | 1.89% |
| **Insurance Class** |  |  |  |
| Blue Cross Blue Shield | 78 | 1863 | 4.02% |
| Commercial | 61 | 1326 | 4.40% |
| Medicare | 48 | 1105 | 4.16% |
| Medicare Managed | 67 | 886 | 7.03% |
| Others | 15 | 240 | 5.88% |
| Medicaid Managed | 21 | 140 | 13.04% |
| **Race** |  |  |  |
| White | 268 | 5088 | 5.00% |
| Others | 9 | 176 | 4.86% |
| Asian | 1 | 128 | 0.78% |
| Hispanic | 11 | 125 | 8.09% |
| Black | 1 | 43 | 2.27% |
| **Language** |  |  |  |
| English | 285 | 5457 | 4.96% |
| Others | 5 | 103 | 4.63% |
| **Charlson Deyo Score** |  |  |  |
| 1-2 | 63 | 1311 | 4.59% |
| 3-4 | 130 | 2766 | 4.49% |
| >=5 | 97 | 1483 | 6.14% |
| **General Adult Score** |  |  |  |
| 0 | 64 | 1751 | 3.53% |
| 1 | 105 | 2251 | 4.46% |
| 2 | 59 | 1006 | 5.54% |
| 3 | 28 | 372 | 7.00% |
| 4 | 19 | 114 | 14.29% |
| 5 | 11 | 35 | 23.91% |
| 6 | 2 | 16 | 11.11% |
| 7 | 1 | 9 | 10.00% |
| 8 | 0 | 4 | 0% |
| 9 | 0 | 2 | 0% |
| 10 | 1 | 0 | 100% |
| **MyDH Active Within Last Year** |  |  |  |
| Yes | 259 | 5204 | 4.74% |
| No | 31 | 356 | 8.01% |
| **Age Group** |  |  |  |
| 50-54 | 63 | 1003 | 5.91% |
| 55-59 | 58 | 1081 | 5.09% |
| 60-64 | 64 | 1236 | 4.92% |
| 65-69 | 61 | 1245 | 4.67% |
| 70-74 | 44 | 995 | 4.23% |
| **State Rank (ADI)** |  |  |  |
| 1 | 7 | 433 | 1.59% |
| 2 | 15 | 410 | 3.53% |
| 3 | 9 | 486 | 1.82% |
| 4 | 17 | 564 | 2.93% |
| 5 | 16 | 775 | 2.02% |
| 6 | 38 | 747 | 4.84% |
| 7 | 52 | 777 | 6.27% |
| 9 | 48 | 512 | 8.57% |
| 8 | 43 | 507 | 7.82% |
| 10 | 45 | 349 | 11.42% |
| **Rural-Urban Commuting Area** |  |  |  |
| 1 | 42 | 1729 | 2.37% |
| 2 | 8 | 295 | 2.64% |
| 3 | 170 | 2143 | 7.35% |
| 4 | 70 | 1392 | 4.79% |

**Table A5:**
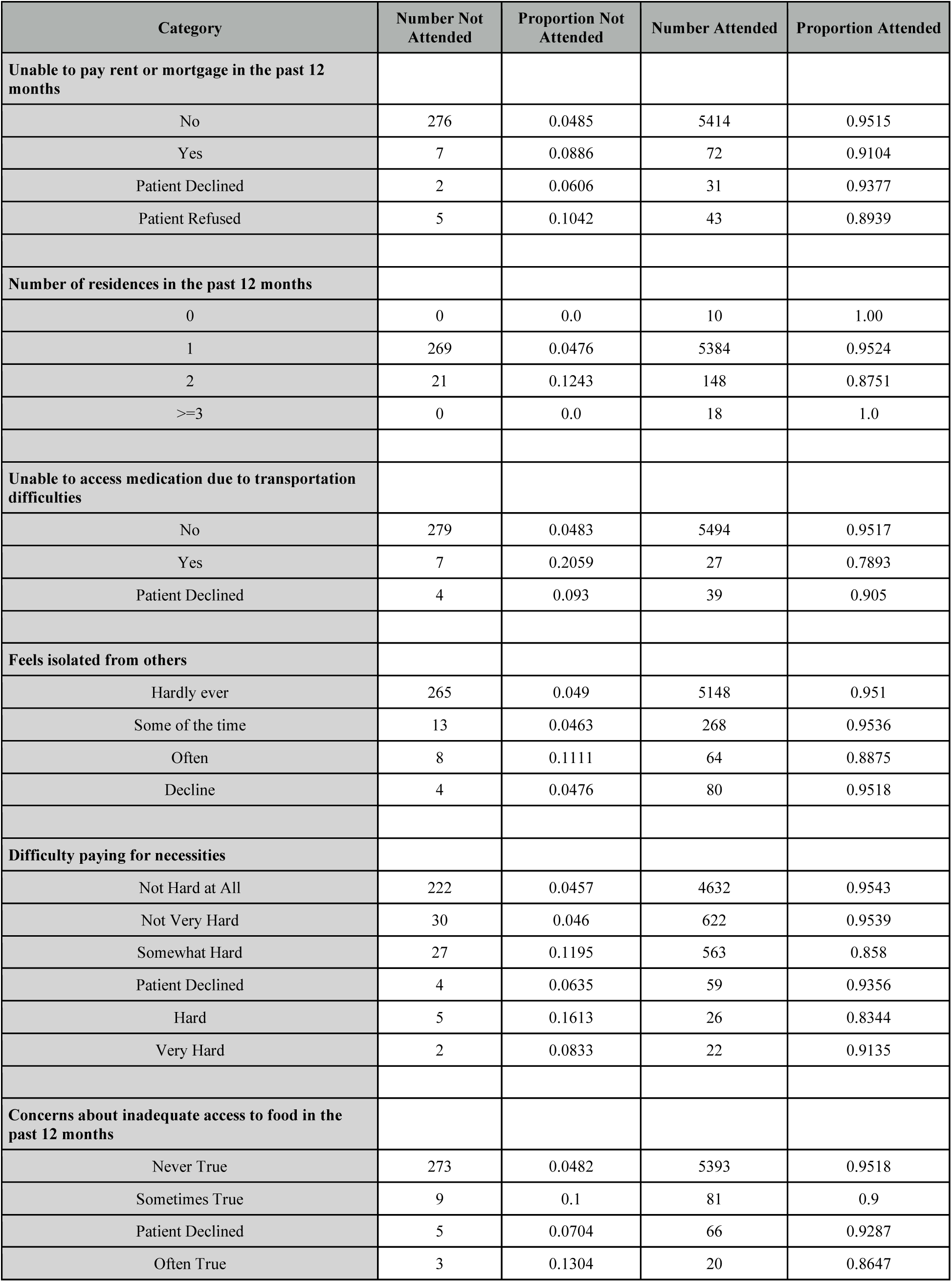
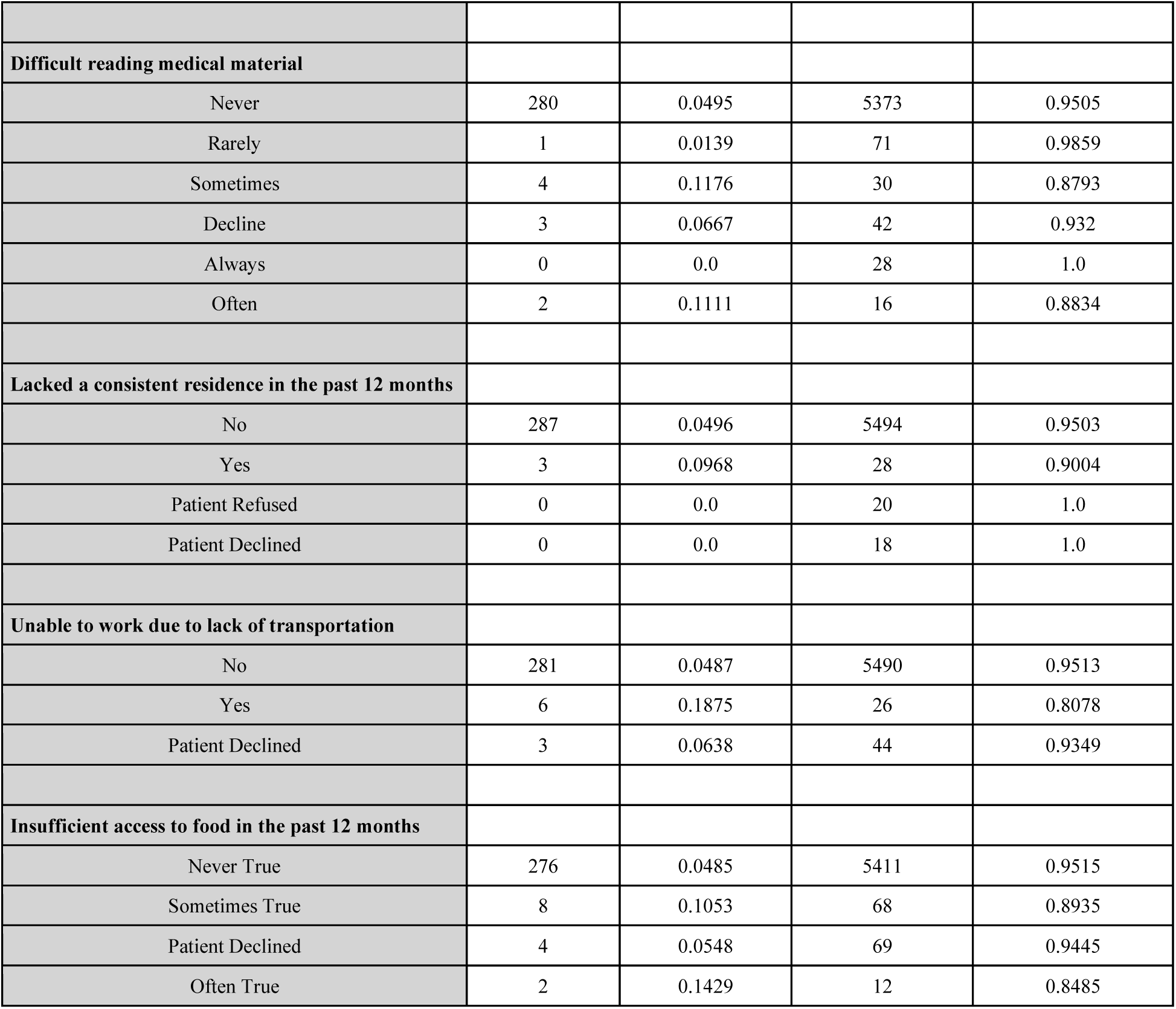
Consolidated Question Counts—Attended.

| Category | Number Not Attended | Proportion Not Attended | Number Attended | Proportion Attended |
| --- | --- | --- | --- | --- |
| **Unable to pay rent or mortgage in the past 12 months** |  |  |  |  |
| No | 276 | 0.0485 | 5414 | 0.9515 |
| Yes | 7 | 0.0886 | 72 | 0.9104 |
| Patient Declined | 2 | 0.0606 | 31 | 0.9377 |
| Patient Refused | 5 | 0.1042 | 43 | 0.8939 |
| **Number of residences in the past 12 months** |  |  |  |  |
| 0 | 0 | 0.0 | 10 | 1.00 |
| 1 | 269 | 0.0476 | 5384 | 0.9524 |
| 2 | 21 | 0.1243 | 148 | 0.8751 |
| >=3 | 0 | 0.0 | 18 | 1.0 |
| **Unable to access medication due to transportation difficulties** |  |  |  |  |
| No | 279 | 0.0483 | 5494 | 0.9517 |
| Yes | 7 | 0.2059 | 27 | 0.7893 |
| Patient Declined | 4 | 0.093 | 39 | 0.905 |
| **Feels isolated from others** |  |  |  |  |
| Hardly ever | 265 | 0.049 | 5148 | 0.951 |
| Some of the time | 13 | 0.0463 | 268 | 0.9536 |
| Often | 8 | 0.1111 | 64 | 0.8875 |
| Decline | 4 | 0.0476 | 80 | 0.9518 |
| **Difficulty paying for necessities** |  |  |  |  |
| Not Hard at All | 222 | 0.0457 | 4632 | 0.9543 |
| Not Very Hard | 30 | 0.046 | 622 | 0.9539 |
| Somewhat Hard | 27 | 0.1195 | 563 | 0.858 |
| Patient Declined | 4 | 0.0635 | 59 | 0.9356 |
| Hard | 5 | 0.1613 | 26 | 0.8344 |
| Very Hard | 2 | 0.0833 | 22 | 0.9135 |
| **Concerns about inadequate access to food in the past 12 months** |  |  |  |  |
| Never True | 273 | 0.0482 | 5393 | 0.9518 |
| Sometimes True | 9 | 0.1 | 81 | 0.9 |
| Patient Declined | 5 | 0.0704 | 66 | 0.9287 |
| Often True | 3 | 0.1304 | 20 | 0.8647 |
| **Difficult reading medical material** |  |  |  |  |
| Never | 280 | 0.0495 | 5373 | 0.9505 |
| Rarely | 1 | 0.0139 | 71 | 0.9859 |
| Sometimes | 4 | 0.1176 | 30 | 0.8793 |
| Decline | 3 | 0.0667 | 42 | 0.932 |
| Always | 0 | 0.0 | 28 | 1.0 |
| Often | 2 | 0.1111 | 16 | 0.8834 |
| **Lacked a consistent residence in the past 12 months** |  |  |  |  |
| No | 287 | 0.0496 | 5494 | 0.9503 |
| Yes | 3 | 0.0968 | 28 | 0.9004 |
| Patient Refused | 0 | 0.0 | 20 | 1.0 |
| Patient Declined | 0 | 0.0 | 18 | 1.0 |
| **Unable to work due to lack of transportation** |  |  |  |  |
| No | 281 | 0.0487 | 5490 | 0.9513 |
| Yes | 6 | 0.1875 | 26 | 0.8078 |
| Patient Declined | 3 | 0.0638 | 44 | 0.9349 |
| **Insufficient access to food in the past 12 months** |  |  |  |  |
| Never True | 276 | 0.0485 | 5411 | 0.9515 |
| Sometimes True | 8 | 0.1053 | 68 | 0.8935 |
| Patient Declined | 4 | 0.0548 | 69 | 0.9445 |
| Often True | 2 | 0.1429 | 12 | 0.8485 |

## Secondary Analyses

Our analytical framework includes several secondary analyses designed to provide deeper insights into factors influencing mammogram screening behaviors. These analyses complement our primary models and offer healthcare systems actionable information for implementing targeted interventions.

## Analysis A1: Scheduling Behavior

### A1.1: SDoH-Only Model

For scheduling behavior, we conducted a systematic exploration at multiple levels. First, we developed a SDoH-only model that isolates SDoH to understand their specific contribution to scheduling behaviors. This targeted approach helped identify which SDoH most significantly influence scheduling uptake, providing healthcare systems with evidence-based insights to design interventions that address the most influential social factors. Our analysis revealed that SDoH variables alone had limited predictive power (test AUC=0.51), with the model’s nested cross validation yielding an average validation AUC of 0.51 and best performance of 0.54. The substantial performance gap between this specialized model and our comprehensive model (AUC=0.71) underscored the complex, inter-connected nature of factors influencing scheduling behavior.

### A1.2: Age-Stratified Analysis

**Table A1.2:** Age-Stratified Model Performance. Area Under the Curve (AUC) values and sample sizes across age groups for previously unseen test sets, demonstrating varying model performance across different age demographics.

| **Age Group** | **Average Validation AUC** | **Test Set AUC** | **Number of Patients in the Test Set** |
| --- | --- | --- | --- |
| 50-54 | 0.72 | 0.68 | 1658 |
| 55-59 | 0.7 | 0.7 | 1845 |
| 60-64 | 0.7 | 0.65 | 1928 |
| 65-69 | 0.71 | 0.67 | 1911 |
| 70-75 | 0.72 | 0.65 | 1550 |

We performed age-stratified evaluations of the light gradient boosting model across five-year intervals (i.e., 50-54, 55-59, 60-64, 65-69, 70-75) to identify how SDoH’s influence varies across demographic groups. This stratification revealed relatively consistent predictive performance across different age groups (Table A1.2). The model showed minimal variation in performance, with average validation AUC values ranging from 0.7 to 0.72 across all age groups. The model performed slightly better for previously-unseen patients aged 50-54 (AUC=0.72, n=1342) and 70-75 (AUC=0.72, n=1212), compared to those aged 55-59 (AUC=0.7, n=1435) and 60-64 (AUC=0.7, n=1619), with the 65-69 age group falling in between (AUC=0.71, n=1505). This consistency suggested that social drivers of scheduling behavior may operate similarly across different life stages, though the slight variations could still be considered when fine-tuning outreach strategies.

### A1.3: Patient-Level Models

To evaluate the independent contribution of individual characteristics to scheduling behavior, we developed a model focusing exclusively on patient-specific factors while excluding system-level variables. This individual-level model concentrated solely on patient characteristics (e.g., age, race, language, insurance class, months since the last mammogram, and ADI) and demonstrated relatively weaker predictive capability (test AUC=0.6) compared to our comprehensive model.

The nested cross validation produced an average validation AUC of 0.65, with the best performance reaching 0.71 across validation folds. These results suggested that individual demographic and socio-economic factors provides limited explanatory power for scheduling behavior.

### A1.4: Clinic-Level Analysis

Our analysis of model performance by individual clinical site revealed substantial variation in predictive capabilities across different organizational settings (Table A1.4). The light gradient boosting machine model demonstrated strong performance in certain facilities, with Clinical Site with 1.8K Patients (average AUC=0.88, test AUC=0.73) and Clinical Site with 6.5K Patients (average AUC=0.74, test AUC=0.69) showing particularly robust predictive power. In contrast, other locations such as Clinical Site with 11K Patients (average AUC=0.66, test AUC=0.63) and Clinical Site with 4K Patients (average AUC=0.67, test AUC=0.67) exhibited modest performance. These facility-level variations in model effectiveness suggested that organizational contexts significantly influence the relationship between SDoH and scheduling behavior, highlighting the importance of organizational factors in implementation success.

**Table A1.4:** Clinical Site-Stratified Model Performance. Area Under the Curve (AUC) values across individual primary care practice locations, demonstrating how model performance varies across different organizational settings within the Dartmouth Health System.

| **Clinical Site (De-identified)** | **Average Validation AUC** | **Test Set AUC** | **Number of Patients in the Test Set** |
| --- | --- | --- | --- |
| Clinical Site with 11k patients | 0.66 | 0.63 | 1807 |
| Clinical Site with 10.7k patients | 0.7 | 0.65 | 1718 |
| Clinical Site with 9.6k patients | 0.69 | 0.65 | 1341 |
| Clinical Site with 6.8k patients | 0.74 | 0.65 | 964 |
| Clinical Site with 6.5k patients | 0.74 | 0.69 | 973 |
| Clinical Site with 4k patients | 0.67 | 0.67 | 697 |
| Clinical Site with 2.2k patients | 0.71 | 0.62 | 352 |
| Clinical Site with 2k patients | 0.68 | 0.57 | 315 |
| Clinical Site with 1.9k patients | 0.69 | 0.66 | 331 |
| Clinical Site with 1.8k patients | 0.88 | 0.73 | 265 |
| Clinical Site with 1k patients | 0.74 | 0.44 | 129 |

### A1.5: Complete Case Analysis

We conducted comparative analyses using complete cases only as a sensitivity check to evaluate how different approaches to missing data management affect implementation insights. The secondary analysis using complete case data demonstrated similar patterns but generally lower predictive performance. The logistic regression and random forest model achieved an average AUC of 0.662 and 0.685 respectively, while the light gradient boosting model showed comparable performance to the imputed data analysis (Average AUC=0.693). This consistency between complete and imputed analyses suggested that our findings are robust to different approaches for handling missing SDoH data, which is an important consideration for healthcare systems implementing SDoH scheduling programs.

The similar pattern of results between complete and imputed analyses, particularly in identifying key predictive factors, reinforced the reliability of our findings for implementation planning. The modest decrease in performance with complete case data suggested that the imputed SDoH data handling method may improve predictive accuracy, though the core relationships between social drivers and scheduling behavior remain stable across analyses.

## Analysis A2: Attendance Behavior

### A2.1: SDoH-Only Attendance Model

We performed a similar SDoH-only analysis on attendance behavior. We developed a model that isolates the SDoH variables to comprehend their specific contribution to screening adherence, in particular the which SDoH most significantly impact screening adherence. Our aim with this analysis was to provide healthcare systems with powerful, evidence-based insights with the hope of creating interventions that address these social factors. Our isolated SDoH analysis, however, revealed that SDoH variables alone have limited predicted power (test AUC = 0.56). The model’s nested cross validation had an average AUC of 0.55 with a best performance of 0.58. The marked performance gap relative to our full model (AUC = 0.69) emphasized the multifaceted and interrelated drivers of scheduling behavior.

### A2.2: Attendance Age Stratified Analysis

For our attendance analysis, we performed age-stratified evaluations of the logistic regression model across five-year intervals (i.e., 50-54, 55-59, 60-64, 65-69, 70-75) to explore how the impact of social determinants of health differs among various demographic groups. Unlike for scheduling behavior, the stratification of the attendance model revealed inconsistent performance across age groups (Table A2.2) and across our testing and validation sets. The model appeared to perform significantly better for patients aged 55-59 (AUC = 0.85, n = 239), than for those 50-54 (AUC = 0.67, n = 183), 60-64 (AUC = 0.68, n = 265), 65-69 (AUC = 0.68, n = 270), 70-75 (AUC = 0.40, n = 213). However, due to the large inconsistencies between the validation and testing performance, it is difficult to make any strong conclusions about the way in which SDoH operate across different life stages. It is important to acknowledge that the small number of patients in each age group may contribute to the model’s inconsistent performance.

**Table A2.2:** Age-Stratified Model Performance. Area Under the Curve (AUC) values and sample sizes across age groups for previously unseen test sets, demonstrating varying model performance across different age demographics.

| Age Group | Average Validation AUC | Test Set AUC | Number of Patients in the Test Set |
| --- | --- | --- | --- |
| 50-54 | 0.77 | 0.67 | 183 |
| 55-59 | 0.75 | 0.85 | 239 |
| 60-64 | 0.69 | 0.68 | 265 |
| 65-69 | 0.68 | 0.68 | 270 |
| 70-75 | 0.69 | 0.40 | 213 |

### A2.3: Attendance Patient Level Model

To assess how individual traits alone influence attendance behavior, we built a model that isolates patient-level factors, deliberately omitting system-related variables. This model included patient characteristics such as age, race, and ADI. The model demonstrated comparable predictive performance to our baseline model (AUC = 0.68). The nested cross validation had an average validation AUC of 0.69. These results suggested that patient-level characteristics alone carry substantial predictive power for attendance behavior, nearly matching the performance of more comprehensive models.

### A2.4: Attendance Clinical Level Analysis

**Table A2.4:** Clinical Site-Stratified Model Performance. Area Under the Curve (AUC) values.

| Clinical Site (De-identified) | Average Validation AUC | Test Set AUC | Number of Patients in the Test Set |
| --- | --- | --- | --- |
| Clinical Site with 11k patients | 0.4 | 0.56 | 263 |
| Clinical Site with 10.7k patients | 0.64 | 0.67 | 252 |
| Clinical Site with 9.6k patients | 0.86 | 0.68 | 235 |
| Clinical Site with 6.8k patients | 0.58 | 0.85 | 87 |
| Clinical Site with 6.5k patients | 0.5 | 0.62 | 114 |
| Clinical Site with 4k patients | 0.35 | 0.75 | 70 |
| Clinical Site with 2.2k patients | 0.23 | 0.82 | 55 |
| Clinical Site with 2k patients | 0.5 | 0.5 | 26 |
| Clinical Sites with 1.9k patients and 1.8k patients | 0.63 | 0.55 | 46 |
| Clinical Site with 1.0k patients | 0.76 | 0.81 | 22 |

For our attendance analysis, we performed clinical site stratified evaluations of the logistic regression model to explore how the impact of SDoH differs across different clinics. Unlike for scheduling behavior, the stratification of the attendance model revealed inconsistent performance across the different clinic groups (Table A2.4) and across our testing and validation sets. Due to the large inconsistencies between the validation and testing performance, it is difficult to make any strong conclusions about the way in which SDoH operate across different clinical site. It is important to acknowledge that the small number of patients in each age group may contribute to the model’s inconsistent performance. Due to this small number of patients, we also combined the results for the clinical site with 1.9k and 1.8k patients based on their geographic similarities.

### A2.5: Attendance Complete Case Analysis

We also conducted a complete case analysis for the attendance model as a sensitivity check. Secondary analysis on our complete data set for our chosen model (elastic-net logistic regression) showed comparable results to our model trained on the imputed dataset. Our logistic regression model trained on the complete dataset achieved an AUC of 0.716 which was comparable to our performance on the imputed dataset. However, we selected to use the imputed dataset because of its larger size to draw more meaningful insights.

